# Exploring social determinants of childhood vaccination based on the National Immunization Surveys Data

**DOI:** 10.1101/2024.10.18.24315767

**Authors:** Felix M. Pabon-Rodriguez

## Abstract

Ensuring children receive vaccinations is essential for their health and community well-being, preventing serious diseases and fostering overall immunity. Nevertheless, the impact of social determinants of health on vaccination access underscores the urgent need for targeted interventions to address disparities and ensure equitable healthcare delivery for all children. This study investigates the impact of several social determinants on childhood vaccination in the United States using data from the National Immunization Surveys from 2010-2022. As a simple approach, ordinal logistic mixed-effect models is used to analyze vaccination patterns among children aged 19– 35 months as reported by parents or guardians. The study findings highlight associations between vaccination and key social factors, including the child’s age group, parents’ or guardians’ education level, and vaccination awareness due to the child being the firstborn. The results from this basic study provide insights into the nuanced relationships that influence vaccination practices.

## 1. INTRODUCTION

Childhood vaccination programs have played a pivotal role in public health by significantly reducing the burden of vaccine-preventable diseases worldwide [1]. Vaccinations during early childhood are crucial for protecting individuals against a myriad of infectious diseases, thereby contributing to the overall well-being of children and communities. Despite the remarkable success of vaccination initiatives, achieving optimal vaccination coverage remains a persistent challenge, particularly due to emerging infectious threats and vaccine hesitancy concerns [2]. Understanding the factors influencing vaccination uptake is essential for devising targeted interventions and policies aimed at promoting equitable access to vaccines and sustaining high vaccination coverage rates [3].

Social determinants of health (SDoH) have garnered increasing attention in recent years as critical factors of health outcomes and healthcare utilization [4,5]. These determinants, encompassing socioeconomic status, education, employment, housing, social support networks, cultural norms, and healthcare access, have effects on vaccination behavior and coverage rates [6,7]. Disparities in vaccination coverage persist among different demographic groups, highlighting the intricate interplay between socioeconomic factors and vaccine accessibility [8]. Furthermore, the impact of SDoH on vaccination decisions extends beyond individual-level factors to encompass broader societal and structural determinants, underscoring the multifaceted nature of vaccination disparities [9,10]. These SDoH influence health behaviors, healthcare utilization patterns, and overall health status, shaping health outcomes across the lifespan [11-13].

By recognizing the impact of SDoH on health outcomes, researchers can develop more comprehensive and holistic approaches to health promotion and disease prevention. Addressing disparities in SDoH not only mitigates health inequities but also enhances the effectiveness of healthcare interventions and health policies [14,15]. Moreover, integrating SDoH into health research enables the identification of modifiable risk factors and the development of targeted interventions to address underlying social determinants, thereby promoting health equity and improving population health outcomes [12,13,16]. Incorporating SDoH into health research also emphasizes the importance of adopting an interdisciplinary and collaborative approach to address complex health challenges. Collaborations between researchers, policymakers, healthcare providers, community organizations, and other stakeholders are essential for implementing multifaceted interventions that target the root causes of health disparities and promote health equity [17,18].

This work presents a simple analysis examining the relationship between SDoH and childhood vaccination using data from the National Immunization Surveys. The study aims to identify potential influences of various SDoH, such as socioeconomic status, education, and geographic location, on the reported number of vaccinations among children aged 19-35 months, and how these associations change over time. An ordinal logistic mixed-effects model is employed to investigate how these social factors affect vaccination, considering the ordinal nature of the vaccination variables (0, 1, 2, 3+ vaccinations). The findings are expected to provide insights into the interplay between social determinants and childhood vaccination patterns, aiming to inform targeted interventions to improve vaccination uptake and address disparities in immunization.

## 2. MATERIALS AND METHODS

### 2.1. Data

The National Immunization Surveys (NIS) for Children (NIS-Child) focuses on children aged 19-35 months residing in the United States, capturing a representative sample of this population for vaccination coverage assessment. The data collected serve as a crucial tool in monitoring the prevalence of recommended vaccinations among 2-year-old children at national, state, and selected local levels, as well as in U.S. territories. The survey encompasses a comprehensive assessment of coverage for key vaccinations, including Diphtheria and Tetanus Toxoids and Acellular Pertussis vaccine (DTaP/DT/DTP), Poliovirus vaccine (Polio), Measles or Measles-Mumps-Rubella vaccine (MMR), Haemophilus Influenzae type b vaccine (Hib), Hepatitis B vaccine (HepB), Varicella Zoster (chickenpox) vaccine (VAR), Pneumococcal Conjugate vaccine (PCV), Rotavirus vaccine (ROT), Hepatitis A vaccine (HepA), and Influenza vaccine (Flu), leading to a total of ten (10) recommended vaccine administration by age 2, where doses vary by schedule. This study investigates the impact of several social determinants on childhood vaccination in the United States using NIS-Child data from 2010-2022.

#### 2.1.1. Social Determinants of Health

While the primary objective of the survey is to assess immunization status, it inherently reflects several social determinants of health (SDoH) that could influence vaccination patterns. Demographic information, including socioeconomic status such as income and education levels, is implicitly embedded in the data and may influence access to healthcare services, including vaccinations. Additionally, the survey’s multi-level approach, collecting data at national, state, and selected local levels, provides insights into potential geographic and regional disparities in vaccination coverage. Furthermore, the inclusion of demographic variables like race and ethnicity offers a glimpse into disparities that may exist among different racial and ethnic groups. Indirect indicators of healthcare access, such as health insurance coverage and the availability of healthcare facilities, can be inferred from the dataset. Moreover, maternal and child health practices, encompassing factors like breastfeeding and prenatal care, may indirectly impact the likelihood of children receiving recommended vaccinations. While the NIS-Child primarily focuses on vaccination coverage, it offers a valuable lens through which researchers can explore the intersection of immunization and various SDoH, contributing to a more comprehensive understanding of public health dynamics among young children, which is exactly the focus of this study.

#### 2.1.2. Health Outcome

For this study, the number of vaccines received by the child is used as the health outcome of interest. It is important to note that this number can vary based on various factors, including parental choices, access to healthcare, and adherence to vaccination schedules. According to data from the NIS, coverage rates for recommended vaccinations among children by age 2 are regularly assessed. Thus, NIS aims to capture the percentage of children who have received specific vaccines according to the recommended schedules. In addition, parents’ reports are a critical component of assessing vaccine coverage, but it is also important to note that the accuracy of these reports can be influenced by factors such as recall bias, understanding of vaccine schedules, and other individual or cultural considerations. Public health initiatives often emphasize the importance of accurate reporting and maintaining up-to-date immunization records. To facilitate the understanding and interpretation of the model being implemented and the results obtained, a categorical variable (with name *vaccination group* and denoted by *V*) is constructed to represent the grouped number of vaccinations reported by parents or guardians. The variable is constructed as shown in Equation 1, where *i* represents a child aged 19-35 months and *t* the survey year.

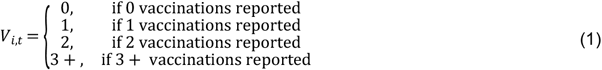

### 2.2. Analysis

The ordinal logistic mixed-effects model expresses the cumulative probability of an ordinal response variable, *V* as previously described, being in or below category *j*. The model is defined as follows (Equation 2).

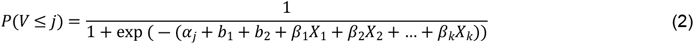

Here, *V* is the ordinal response variable, *X*_1_,*X*_2_,…,*X*_*k*_ are the SDoH of interest, *α*_*j*_ is the intercept of the cumulative log-odds at category *j, b*_1_ is the random effect for census region, *b*_2_ a random effect for year, and *β*_1_,*β*_2_,…,*β*_*k*_ are the estimated coefficients for the SDoH of interest. This model estimates the cumulative probability of being in or below category *j* based on the values of the SDoH of interest. The logistic function 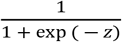, shown in the right-hand side of the previous equation, is used to transform the linear combination of predictors into a probability. The coefficients *β*_1_,*β*_2_,…,*β*_*k*_ represent the impact of each predictor variable on the log-odds of the ordinal response variable. A positive coefficient indicates an increase in the log-odds, while a negative coefficient indicates a decrease. The intercepts *α*_*j*_ represent the cumulative log odds at the baseline level (category 0) and each subsequent category. They determine the starting point of the cumulative probabilities for each category. This model can be applied to investigate the influence of predictor variables on the probabilities of different categories. The model can also be written in its log-odds format as shown in Equation 3.

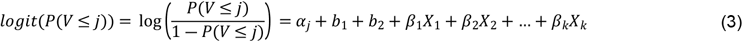

## 3. RESULTS

The goal of this study lies primarily in the assessment of the impact of several the social determinants of health on vaccination coverage by looking at their statistical significance and how this impact has changed over time from 2010 to 2022. Tables 1 provides the definition of the SDoH of interest for this study. To help readers with knowledge with the NIS-Child datasets, this table uses the same names (labels) as described on the website. Figure 1 shows the number of children being vaccinated over time, where the 30-35 months category always shows greater coverage compared to the other groups. This graph also shows that coverage trajectories are very similar for the first two age groups.

**Figure 1.**
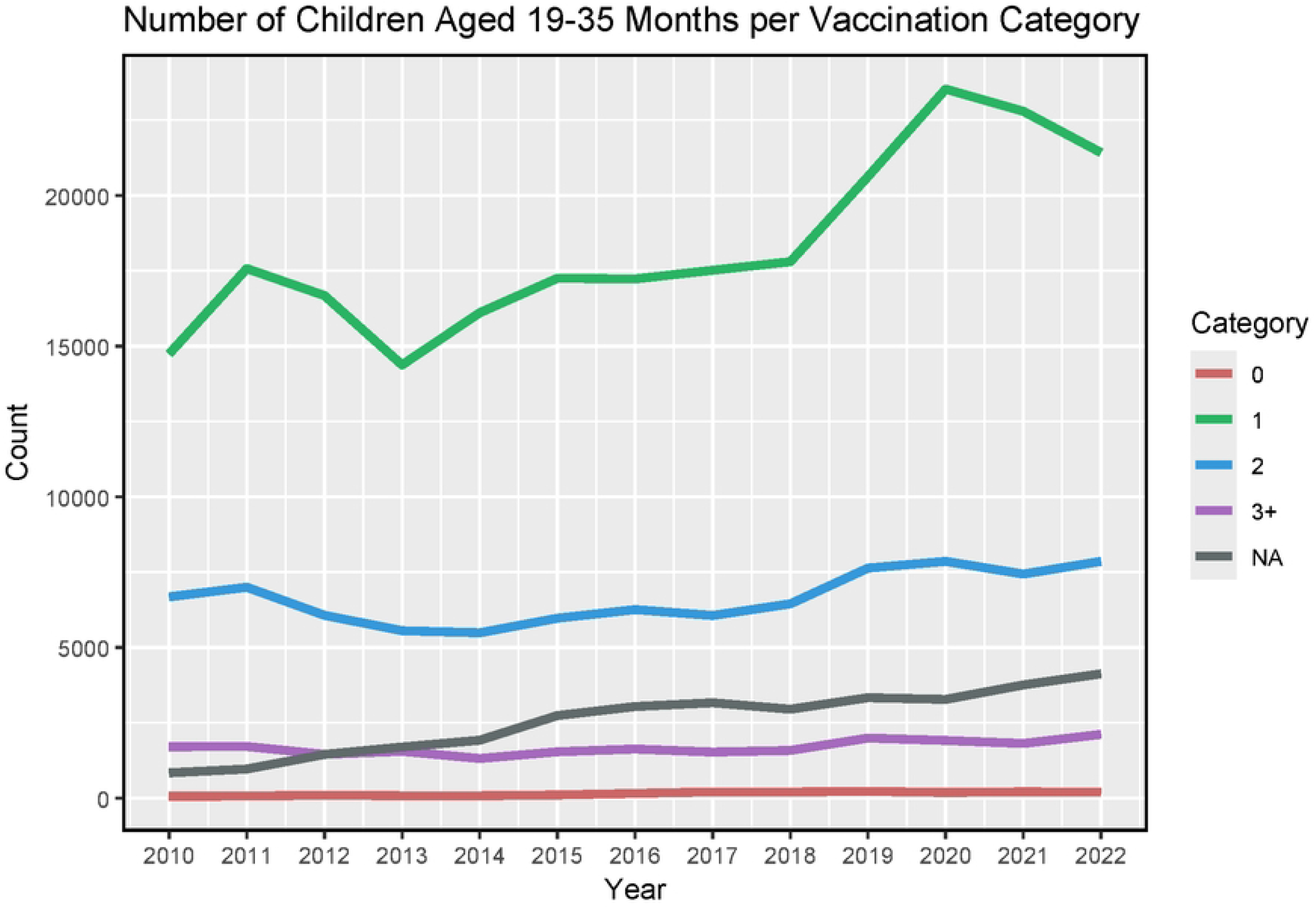
Number of children per year in each age group over the 2010-2022 period.

If we look at the census region (Figure 2), the number of 3+ vaccinations is lower compared to other categories. The South region shows the highest number of vaccinated children with 1 or 2 types. Missing values are affecting these trajectories, as shown in the last plot of Figure 2. When looking at the Hispanic origins of the children (Figure 3), more children from non-Hispanic origins showed higher numbers of vaccines from 2010 to 2022. The poverty-to-income ratio is the ratio between the total income of the children’s family and the poverty threshold. A ratio of less than 1 means the income is below the poverty level, 1 means the income and poverty level are the same, and greater than 1 means the income is above the poverty level. From the data, we found that poverty-to-income ratio patterns look similar over time (Figure 4), with a higher density above the poverty level, but a noticeable peak below 1 is of concern as well.

**Table 1.** Definition of the SDoH variables of interest.

| Variable | Definition |
| --- | --- |
| D6R | Number of vaccination providers identified by respondent |
| YEAR | Year of interview |
| CEN_REG | Census region based on true state of residence |
| AGEGRP | Age category of children (19-23, 24-29, 30-35 months) |
| C1R | Number of people in household |
| CBF_01 | Was the child ever breastfed or fed breast milk? |
| CWIC_02 | Is the child currently receiving WIC benefits? |
| EDUC1 | Education of mother categories |
| FRSTBRN | First born status of child |
| I_HISP_K | Hispanic origin of child |
| INCPORAR | Income to poverty ratio |
| LANGUAGE | Language in which interview was conducted |
| M_AGEGRP | Age of mother categories |
| MARITAL2 | Marital status of mother |
| RACE_K | Race of child |
| RACEETHK | Race/ethnicity of child |
| SEX | Gender of child |
| INS_STAT_I | Was the child not covered by any health insurance at any time? |

**Figure 2.**
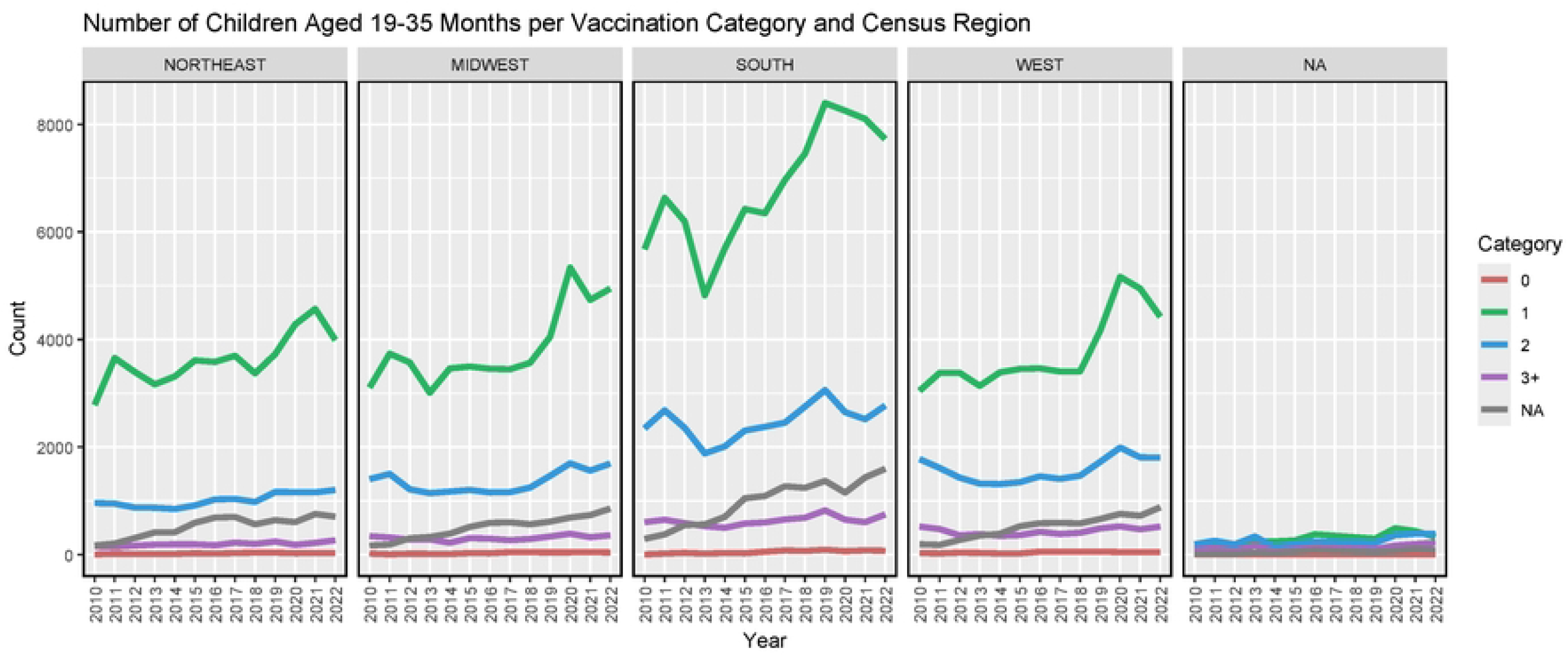
Number of children per year at each vaccination group and census region over the 2010-2022 period. The ‘NA’ plotting area is used to allocate children with missing census regions, and the ‘NA’ in the legend corresponds to missing information about vaccination. The joint ‘NA-NA’ pattern in the last plot correspond to counts with both missing census region and missing vaccination information.

**Figure 3.**
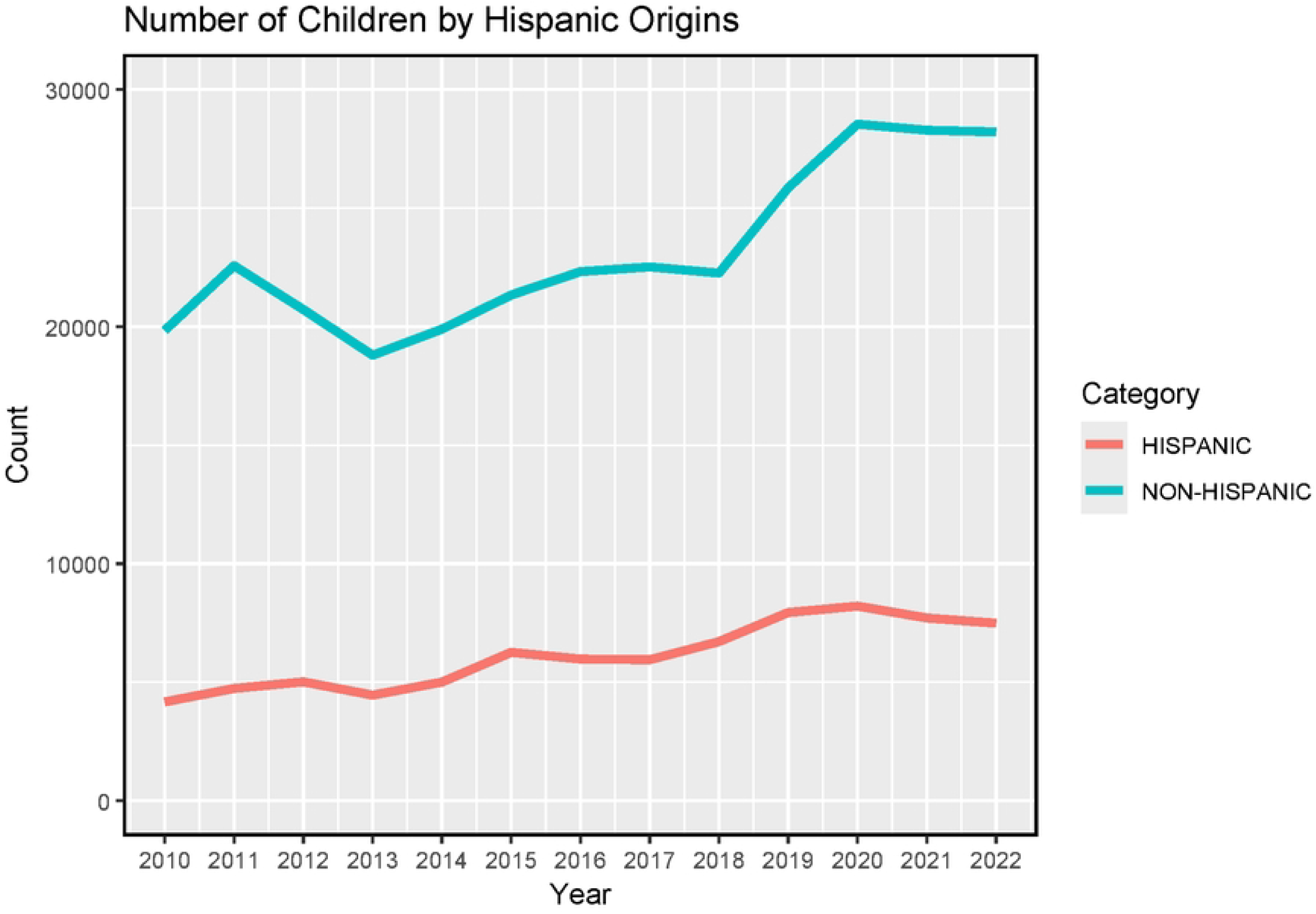
Number of children from Hispanic origins that have received vaccinations.

**Figure 4.**
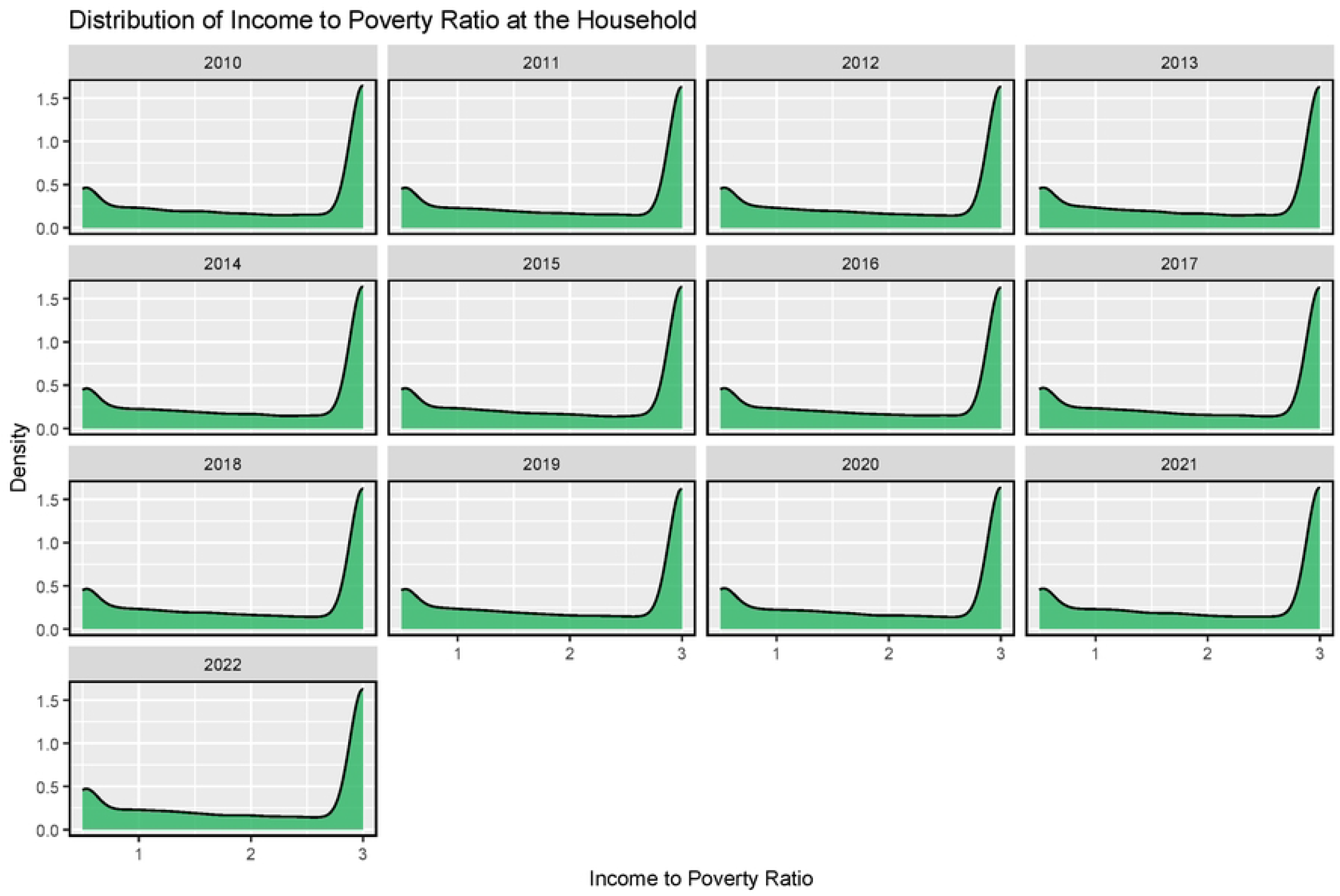
Income to Poverty Ratio per Year. The poverty-to-income ratio is the ratio of a family’s total income divided by its poverty threshold. A ratio of less than 1 means the income is below the poverty level, 1 means the income and poverty level are the same, and greater than 1 means the income is above the poverty level.

Figure 5 illustrates the number of children covered by health insurance. From this plot, we see that the number of missing information regarding health insurance coverage has increased dramatically since 2010. While the number of children covered by health insurance is steadily high compared to the group of children not covered by health insurance, the issue of missing information is worrisome. To explore the initial associations between each considered SDoH and childhood vaccination coverage, we also conducted a Fisher exact test to assess the significance of these associations via contingency tables formed between the outcome and the factor without considering the other variables. From this, the study suggests that in each year from 2010 to 2022, the age group and Hispanic origins of the children, education level, and language of the parent or guardian, and if the child is firstborn, are statistically associated with the vaccination status at the 5% significance level.

**Figure 5.**
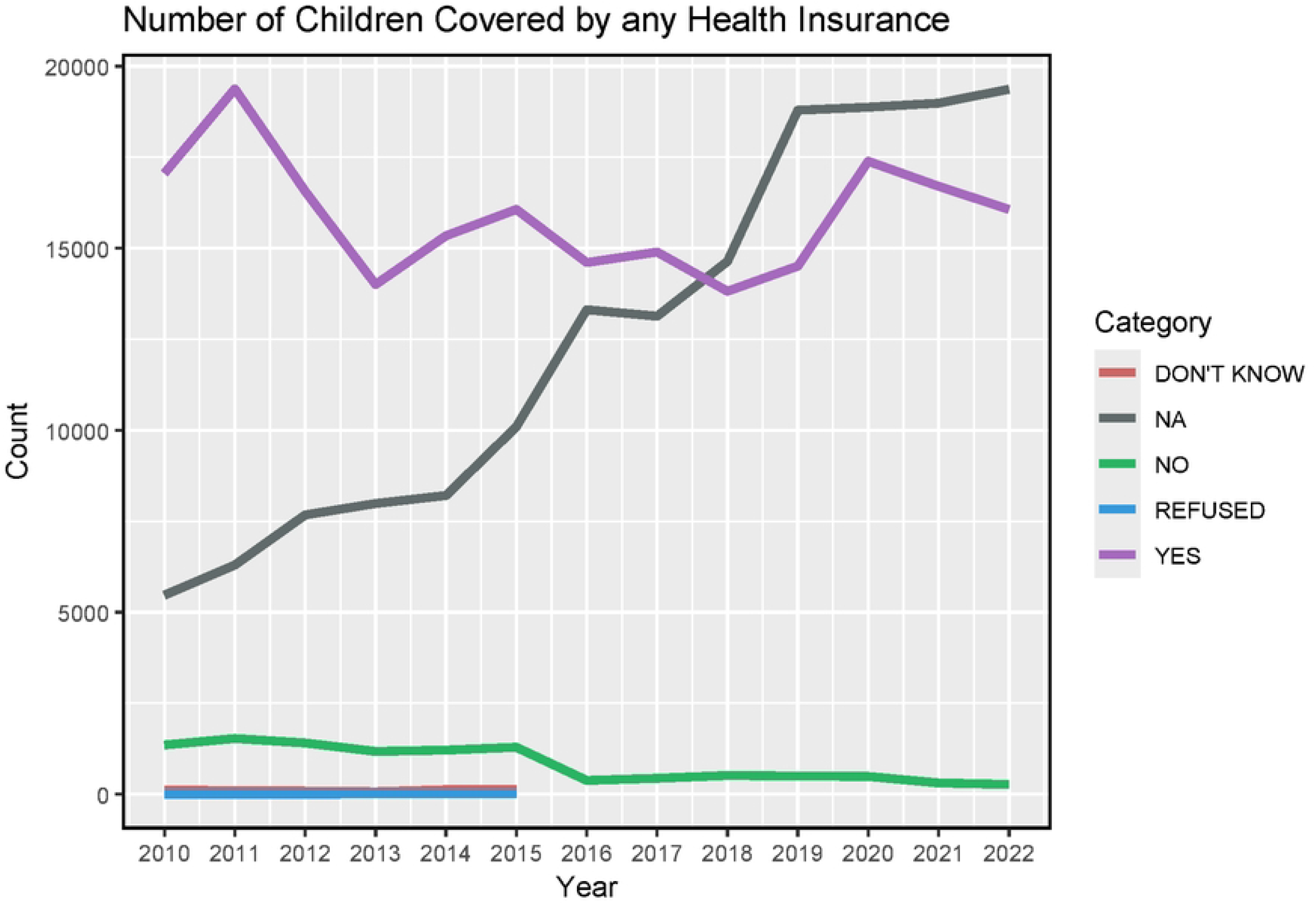
Number of children from covered by any health insurance.

### 3.1. Year-specific models

The first analysis uses Equation 1 with all covariates of interests and a random effect for census region. This model revealed some significant associations between SDoH variables and childhood vaccination. Figure 6 illustrates the statistical significance over time of various factors with the vaccination group. The red and blue colors represent if the odds ratios are greater than 1 or not (red = no, blue = yes). In the x-axis, we look at the significance level of the predictor, with names shown in the y-axis. Value 0 means it is not significant, while 1 means it is statistically significant. Several SDoH factors significantly influence childhood vaccination behavior. Specifically, the child’s age group, parents’ or guardians’ education level, vaccination awareness due to the child being the firstborn, and the income-to-poverty ratio were found to be significant predictors of vaccination coverage among children aged 19-35 months. Overall, these findings underscore the importance of addressing social determinants such as education and awareness programs in promoting childhood vaccination and reducing disparities in immunization rates.

**Figure 6.**
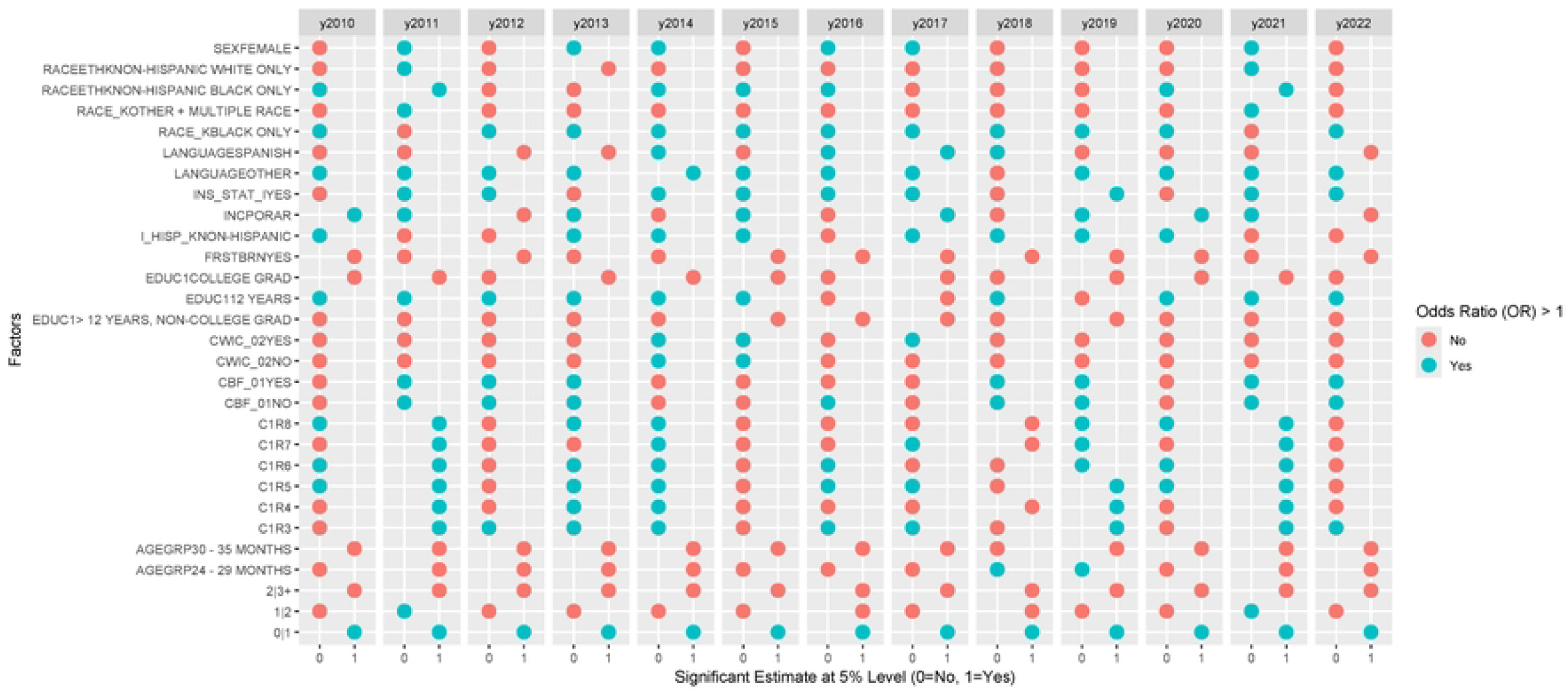
Statistical summary of the year-specific model. The red and blue colors represent whether the odds ratios are greater than 1 or not (red = no, blue = yes). In the x-axis, we look at the significance level of the predictor, with names shown in the y-axis. Value 0 means the factor is not significant, while 1 means it is statistically significant.

### 3.2. Full ordinal logistic mixed-effects model

In this second model, both random effects for census region and year were included. Among factors that were flagged as positively associated with increased vaccination, we have both older age groups (24-29 months, 30-35 months) of the children, the group of children who were breastfed or fed with breast milk, higher education in the mothers, first born child, and Spanish-speaking mothers. All these factors were statistically associated with increased vaccination group membership at the 5% level. On the other hand, among the SDoH factors associated with a decreased vaccination, we have children with Hispanic origin, higher income to poverty ratio, group of mothers older than 30 years old, mother who lives with a partner, children from the Hispanic Black group, indicator that children were not covered by any health insurance. All these factors were statistically associated with a decreased vaccination group membership at the 5% level. Figure 7 summarize these results.

**Figure 7.**
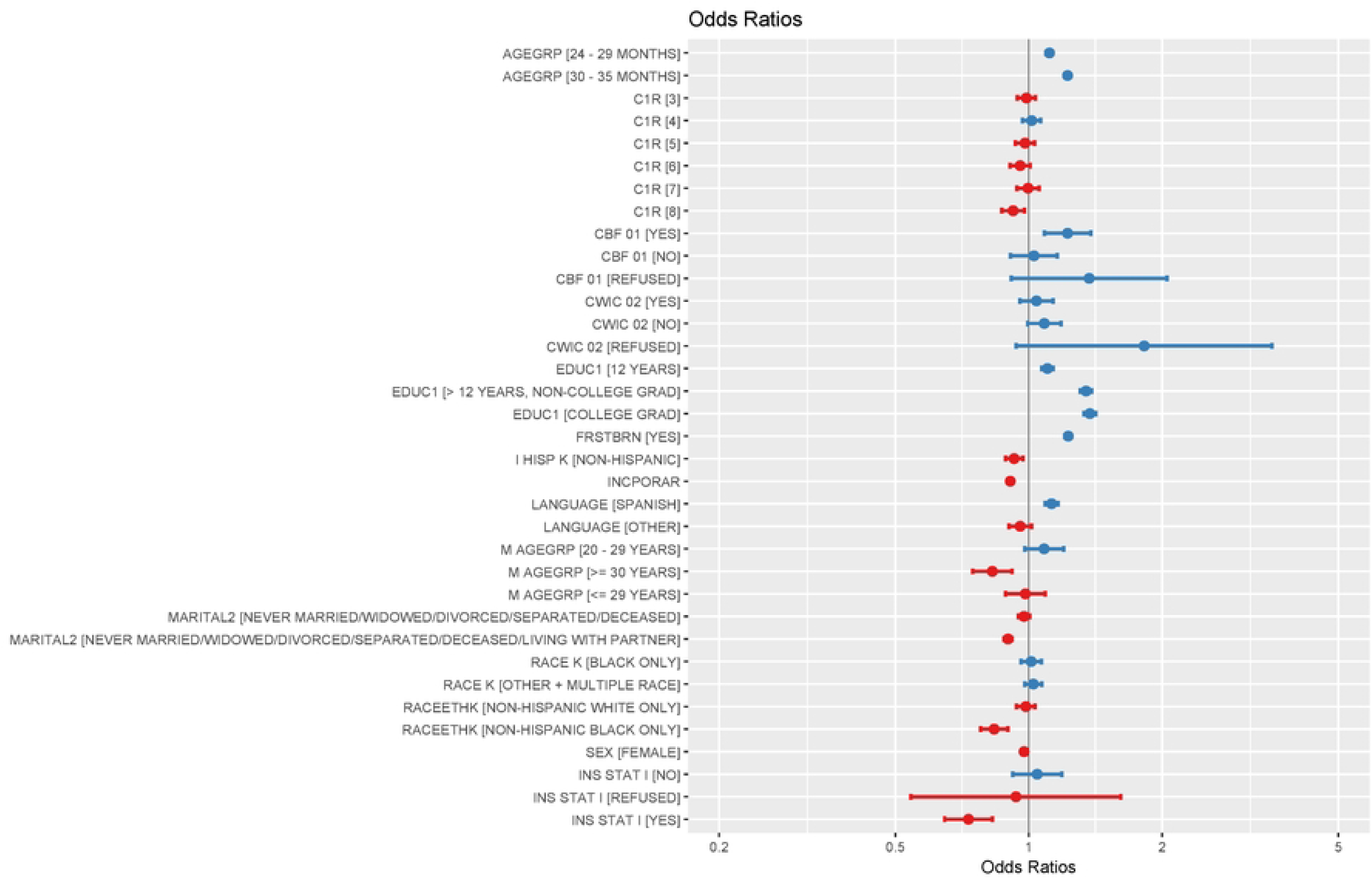
Statistical summary of the full ordinal logistic mixed effects model. The red and blue colors represent whether the odds ratios are greater than 1 or not (red = no, blue = yes).

## 4. DISCUSSION

Moving forward, future research endeavors could expand upon the findings of this study to delve deeper into the mechanisms underlying the observed associations between social determinants of health and childhood vaccination. One avenue for exploration could involve longitudinal studies to track vaccination patterns over time and assess the long-term impact of SDoH on immunization rates. Additionally, qualitative research methods, such as interviews or focus groups with parents and healthcare providers, could provide valuable insights into the barriers and facilitators to vaccination uptake within different demographic groups.

Building on the identified disparities in vaccination, potential initiative implementations should aim to address the root causes of these disparities and promote health equity. Community-based interventions, such as mobile vaccination clinics or outreach programs targeting underserved populations, could improve access to vaccinations in areas with low coverage rates. Furthermore, educational campaigns tailored to specific demographic groups could enhance vaccine awareness and address misconceptions about vaccination safety and efficacy.

Incorporating technology and digital health solutions may also play a crucial role in improving vaccination coverage. Tools such as reminder systems, telehealth consultations, and mobile applications for vaccine scheduling and information dissemination could help overcome logistical barriers and increase vaccine uptake among children and their families. Collaborative efforts between healthcare providers, public health agencies, community organizations, and policymakers will be essential for implementing and evaluating the effectiveness of these initiatives. Overall, future research and initiatives should prioritize strategies that promote equitable access to vaccinations while addressing the social, economic, and cultural factors that contribute to disparities in vaccination coverage. By addressing these challenges, we can work towards achieving higher vaccination rates and better overall health outcomes for children and communities.

## 5. CONCLUSION

This study reveals key social determinants impacting childhood vaccination, identifying factors like age groups, breastfeeding, maternal education, and economic conditions as crucial influences. The disparities observed underscore the urgency of developing strategies that specifically address the unique challenges encountered by vulnerable groups, including children from Hispanic and/or Black groups, uninsured families, and those experiencing economic hardship. Future initiatives should focus on enhancing accessibility through innovative approaches such as community-driven programs, digital health solutions, and culturally sensitive outreach, especially to boost vaccination coverage, ultimately advancing health equity and improving vaccination outcomes for all children.

## 7. ETHICS STATEMENT

The Indiana University Human Research Protection Program (HRPP) staff determined the analysis done in this study is not human subject research and, therefore, did not require further Institutional Review Board (IRB) review before conducting the study.

## 8. DATA AVAILABILITY STATEMENT

The data used for this study is sourced from the National Immunization Surveys database, available on the Centers for Disease Control and Prevention (CDC) website. Access to the dataset can be obtained from the CDC’s official data repository, ensuring transparency and reproducibility in research findings.

## REFERENCES

[1] Andre FE, Booy R, Bock HL, Clemens J, Datta SK, John TJ, Lee BW, Lolekha S, Peltola H, Ruff TA, Santosham M. Vaccination greatly reduces disease, disability, death and inequity worldwide. Bulletin of the World health organization. 2008;86:140–6.

[2] Dubé E, Vivion M, MacDonald NE. Vaccine hesitancy, vaccine refusal and the anti-vaccine movement: influence, impact and implications. Expert review of vaccines. 2015 Jan 2;14(1):99–117.

[3] Opel DJ, Taylor JA, Mangione-Smith R, Solomon C, Zhao C, Catz S, Martin D. Validity and reliability of a survey to identify vaccine-hesitant parents. Vaccine. 2011 Sep 2;29(38):6598–605.

[4] WHO Commission on Social Determinants of Health, World Health Organization. Closing the gap in a generation: health equity through action on the social determinants of health: Commission on Social Determinants of Health final report. World Health Organization; 2008.

[5] Solar O, Irwin A. A conceptual framework for action on the social determinants of health. WHO Document Production Services; 2010.

[6] Luman ET, Barker LE, Shaw KM, McCauley MM, Buehler JW, Pickering LK. Timeliness of childhood vaccinations in the United States: days undervaccinated and number of vaccines delayed. Jama. 2005 Mar 9;293(10):1204–11.

[7] Larson HJ, Jarrett C, Eckersberger E, Smith DM, Paterson P. Understanding vaccine hesitancy around vaccines and vaccination from a global perspective: a systematic review of published literature, 2007–2012. Vaccine. 2014 Apr 17;32(19):2150–9.

[8] McDonald HI, Tessier E, White JM, Woodruff M, Knowles C, Bates C, Parry J, Walker JL, Scott JA, Smeeth L, Yarwood J. Early impact of the coronavirus disease (COVID-19) pandemic and physical distancing measures on routine childhood vaccinations in England, January to April 2020. Eurosurveillance. 2020 May 14;25(19):2000848.

[9] Phadke VK, Bednarczyk RA, Salmon DA, Omer SB. Association between vaccine refusal and vaccine-preventable diseases in the United States: a review of measles and pertussis. Jama. 2016 Mar 15;315(11):1149–58.

[10] Olusanya OA, Bednarczyk RA, Davis RL, Shaban-Nejad A. Addressing parental vaccine hesitancy and other barriers to childhood/adolescent vaccination uptake during the coronavirus (COVID-19) pandemic. Frontiers in immunology. 2021 Mar 18;12:663074.

[11] P. Braveman, S. Egerter, and D. R. Williams, “The social determinants of health: coming of age,” Annu Rev Public Health, vol. 32, pp. 381–398, 2011.

[12] Link BG, Phelan J. Social conditions as fundamental causes of disease. Journal of health and social behavior. 1995 Jan 1:80–94.

[13] Phelan JC, Link BG, Tehranifar P. Social conditions as fundamental causes of health inequalities: theory, evidence, and policy implications. Journal of health and social behavior. 2010 Mar;51(1_suppl):S28–40.

[14] Graham H. Social determinants and their unequal distribution: clarifying policy understandings. The Milbank Quarterly. 2004 Mar;82(1):101–24.

[15] Adler NE, Stewart J. Health disparities across the lifespan: meaning, methods, and mechanisms. Annals of the New York Academy of Sciences. 2010 Feb;1186(1):5–23.

[16] Berkman LF, Kawachi I, Glymour MM, editors. Social epidemiology. Oxford University Press; 2014.

[17] Minkler M, Blackwell AG, Thompson M, Tamir H. Community-based participatory research: implications for public health funding. American journal of public health. 2003 Aug;93(8):1210–3.

[18] Diez Roux AV. Complex systems thinking and current impasses in health disparities research. American journal of public health. 2011 Sep;101(9):1627–34.

